# Health-care workers in gastrointestinal endoscopy are at higher risk for SARS-CoV-2 infection compared to other aerosol-generating disciplines

**DOI:** 10.1101/2021.09.20.21263566

**Authors:** Christoph Römmele, Alanna Ebigbo, Maria Kahn, Stephan Zellmer, Anna Muzalyova, Gertrud Hammel, Christina Bartenschlager, Albert Beyer, Jonas Rosendahl, Tilo Schlittenbauer, Johannes Zenk, Bilal Al-Nawas, Roland Frankenberger, Jürgen Hoffmann, Christoph Arens, Frank Lammert, Claudia Traidl-Hoffmann, Helmut Messmann

## Abstract

**Objective:** Healthcare workers (HCW) are at high risk of SARS-CoV-2 infection due to exposure to potentially infectious material, especially during aerosol-generating procedures (AGP). We aimed to investigate the prevalence of infection among HCW in medical disciplines with AGP.

**Design:** A nationwide questionnaire-based study in in- and outpatient settings was conducted between 12/16/2020 and 01/24/2021. Data on SARS-CoV-2 infections among HCW and potential risk factors were investigated.

**Results:** 2,070 healthcare facilities with 25,113 employees were included in the study. Despite a higher rate of pre-interventional testing, clinics treated three times more confirmed SARS-CoV-2 cases than private practices (28.8% vs. 88.4%, p<0.001). Overall infection rate among HCW accounted for 4.7%. Multivariate analysis revealed that ZIP-regions having comparably higher incidences were significantly associated with increased risk of infection. Furthermore, clinical setting and the GIE specialty have more than double the risk of infection (OR 2.63; 95% CI 2.501-2.817, p<0.01 and OR 2.35; 95% CI 2.245-2.498, p<0.01). The number of procedures performed per day was also significantly associated with an increased risk of infection (OR 1.01; 95% CI 1.007-1.014), p<0.01). No treatment of confirmed SARS-CoV-2 cases was tending to lower the risk of infection (OR 0.72; 95% CI 0.507-1.025, p=0.068).

**Conclusion:** HCW in GIE seem to be at higher risk of infection than those in other AGP, especially in the clinical setting. Regions having comparably higher incidences as well as the number of procedures performed per day were also significantly associated with increased risk of infection.

**Significance of this study:** *What is already known on this subject?:* Health care workers, especially those exposed to aerosol generating procedures, are assumed to have an increased risk of SARS-CoV-2 infection. However, data confirming this are lacking, especially for the outpatient care setting.

*What are the new findings?:* Health care workers in gastrointestinal endoscopy have a higher risk of SARS-CoV-2-infection than in other AGPs. This risk is particularly higher - in clinical settings compared to private practices
- in regions having comparably higher incidences
- the more procedures are performed per day

*How might it impact on clinical practice in the foreseeable future?:* Our study suggests making additional efforts to protect HCW in the gastrointestinal work field.

## INTRODUCTION

For more than a year, the “coronavirus disease 2019” (COVID-19) has kept the world and especially the health care sector on tenterhooks. A small outbreak of the virus “severe acute respiratory syndrome coronavirus type 2” (SARS-CoV-2) has since developed into a worldwide pandemic with over 200 million cases (as of 08/30/2021),[1].

Health care workers (HCW) have been particularly exposed during the pandemic, and data shows an increased infection rate among HCW compared to the general population,[2]. Data from different countries emphasise the increased risk for HCW, especially for those with direct patient contact,[3, 4]. Based on these data and the risk of transmission between HCW, the Standing Committee on Vaccination (STIKO) issued a prioritized vaccination recommendation for people working in medical facilities. Transmission of SARS-CoV-2 mainly takes place via respiratory droplets as well as aerosol,[5]. Numerous medical procedures typical for specific medical disciplines are widely recognized to generate aerosols and therefore are supposed to increase the risk of infection. Therefore, HCW who carry out aerosol-generating procedures (AGP) or activities close to patients’ faces were given higher priority for vaccination in Germany, even though real-world data demonstrating the increased risk is limited,[5]. In particular, evidence for this within the outpatient care sector is lacking.

As a part of the collaborative project B-FAST of the Network of University Medicine (NUM), initiated by the German Federal Ministry of Education and Research, Augsburg University Hospital was commissioned to acquire data on facial, and AGP-associated medical subspecialties such as gastrointestinal endoscopy (GIE), otorhinolaryngology (ORL), oral and maxillofacial surgery (OMS) and dentistry. The study was supported by the Bavarian State Ministry of Science and Arts, as well as the respective professional societies, including the German Society of Gastroenterology, Digestive and Metabolic Diseases (DGVS), the German Society of Dentistry and Oral Medicine (DGZMK), the German Society of Oral and Maxillofacial Surgery (DGMKG), The German Society of Oto-Rhino-Laryngology, Head and Neck Surgery (DGHNO-KHC) and the Professional Association of Gastroenterologists in Private Practice (bng).

The study’s objective was to investigate the prevalence of SARS-CoV-2 infection among HCW exposed to AGP in in- and outpatient settings, the suspected source of infection, and identify potential risk or protective factors for infections in the specialties mentioned above.

## MATERIAL AND METHODS

### Questionnaire

The present study is a descriptive, explorative, cross-sectional, questionnaire-based study conducted in Germany between the 16^th^ of December and the 24^th^ of January.

The questionnaire for the survey was designed based on detailed literature research and on expert suggestions provided by the respective disciplines GIE, ORL, OMS, and dentistry (Supplement 1). Besides descriptive data such as healthcare delivery setting and medical specialty, the questionnaire focused on the prevalence of SARS-CoV-2 infection, presumed source of infection, treatment of confirmed SARS-CoV-2 cases, and pre-interventional testing of patients. The first two digits of the ZIP-code of each participating medical facility were inquired to assign medical facilities to one of ten ZIP-code regions in order to correlate infection rate among HCW and incidence in the region.

The target group of the present questionnaire was inpatient and outpatient medical facilities of the four specialties of research interest with a particular focus on GIE. Eligible participants were defined as healthcare delivery facilities attributed to four medical disciplines such as GIE, ORL, OMS, or dentistry. Study participants were recruited via e-mail distribution of the respective professional societies (DGVS, DGZMK, DGMKG, DGHNO-KHC, bng). The heads of department or private practice owners were contacted via e-mail and encouraged to fill in an online questionnaire implemented in UniPark©. Participation in the survey was anonymous and completely voluntary, with no direct contact to the study site.

### Statistical analysis

The statistical analysis was performed using SPSS version 27.0. The categorical variables such as ZIP-code region, medical specialty, type of medical facility, the presumed source of infection, pre-interventional testing of patients, and treatment of confirmed SARS-CoV-2 cases are presented as absolute frequencies and percentages. The interval-scaled variables such as the number of employees and the number of procedures performed per day are presented as mean values and standard deviations. HCW status was calculated as the proportion of the SARS-Cov-2 positive infections among HCW to the underlying total population and expressed as percentages. Covid-19 incidences were calculated using official county-granular data from the Robert Koch Institute (RKI), the leading governmental institution in biomedicine in Germany, aggregated to 10 ZIP-code regions,[6].

In this scientific manuscript, the GIE data are compared with the aggregated data of the disciplines ORL, OMS, and dentistry denoted as Non-GIE. When appropriate, the relationships between nominal-scaled variables were tested inferentially using Chi-square independence tests or Fisher’s exact test. The comparison of GIE and Non-GIE facilities regarding the presumed source of infection was adjusted with Cochran-Mantel-Haenszel-Test to control the confounding effect of the facility healthcare delivery setting. Mean values were compared using Mann-Whitney-U test. The analysis of the risk factors associated with infections among HCW was carried out using multivariate logistic regression with the occurrence of a SARS-CoV-2 infection as a dependent variable and potential influencing variables considered in the manuscript as independent variables. The significance level was set as p < 0.05.

### Patient and public involvement

As a part of the collaborative project B-FAST of the Network of University Medicine (NUM), initiated by the German Federal Ministry of Education and Research, Augsburg University Hospital was commissioned to acquire data on facial, and AGP-associated medical subspecialties. No patients or the public were asked for advice on interpretation or writing up the results.

## RESULTS

### Sample characteristics

Two thousand ninety-six of the more than 20,000 contacted facilities participated in the survey. Twenty-six facilities were excluded from the data analysis based on prespecified eligibility criteria. Consequently, 2,070 remaining questionnaires were analysed, of which 113 (5.5%) had non-exclusionary missing data. Of the 2070 facilities, 1,828 (88.3%) were private practices and 242 (11.7%) were clinics/hospitals. Among included facilities, GIE private practices accounted for 284 (13.7%) facilities, whereas GIE clinics were represented by 145 (7.0%) hospitals (Table 1). The distribution of the Non-GIE facilities between the different disciplines can be found in the supplement (Supplement 1)

**Table 1:**
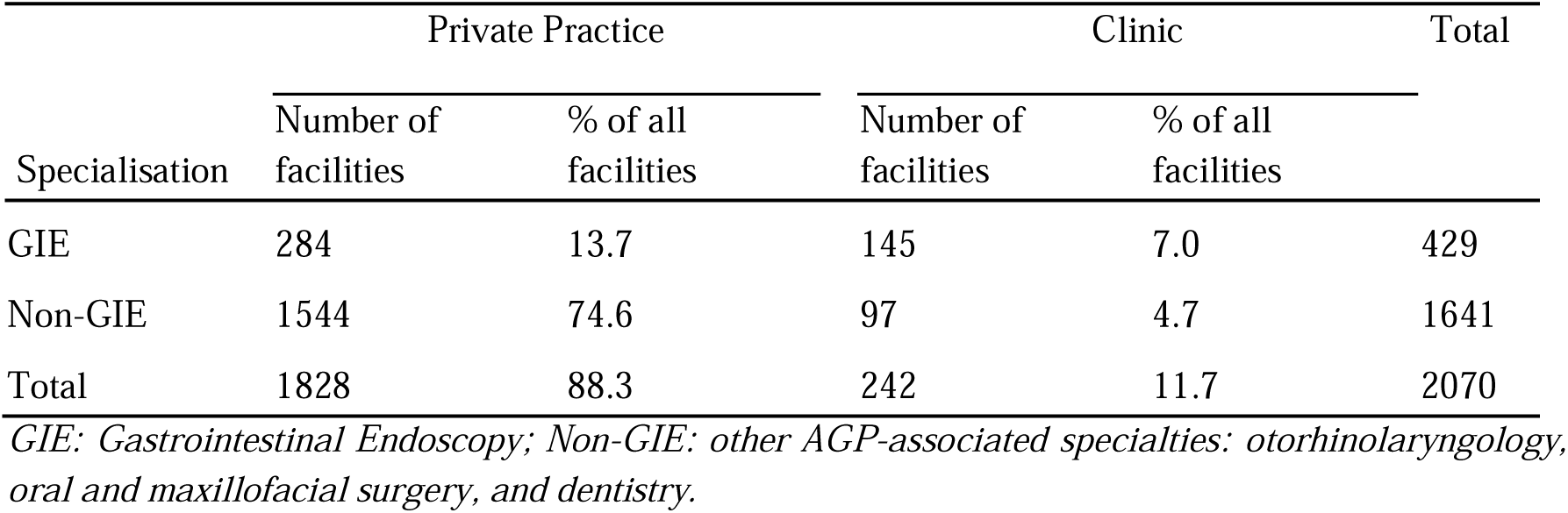
Absolute and percentage distribution of cases by type of institution and specialty

| Specialisation | Private Practice |  | Clinic |  | Total |
| --- | --- | --- | --- | --- | --- |
|  | Number of facilities | % of all facilities | Number of facilities | % of all facilities |  |
| GIE | 284 | 13.7 | 145 | 7.0 | 429 |
| Non-GIE | 1544 | 74.6 | 97 | 4.7 | 1641 |
| Total | 1828 | 88.3 | 242 | 11.7 | 2070 |

*GIE: Gastrointestinal Endoscopy; Non-GIE: other AGP-associated specialties: otorhinolaryngology, oral and maxillofacial surgery, and dentistry.*

The most significant share of participating medical facilities belonged to the districts with the ZIP codes 80-89 (13.4%), and the most seldomly represented district were those of 01-09 (4.8%) (Supplemental 2). Regarding the rate of new infections, in the period from 40^th^ calendar week 2020 (beginning of the second COVID-19 wave) to third calendar week 2021 (the end of the survey) the highest mean incidence was observed in the ZIP-regions 01-09 with 195 (SD=143.0) followed by 90-99 with 147 (SD=90.3) and 80-89 with 143 (SD=71.7) located in the eastern part of the country. The lowest mean incidences were reported in ZIP-regions 20-29 with 72(SD=38.2) in the northern and 30-39 with 108(SD=60.6) central part of Germany (Figure 1).

**Figure 1:**
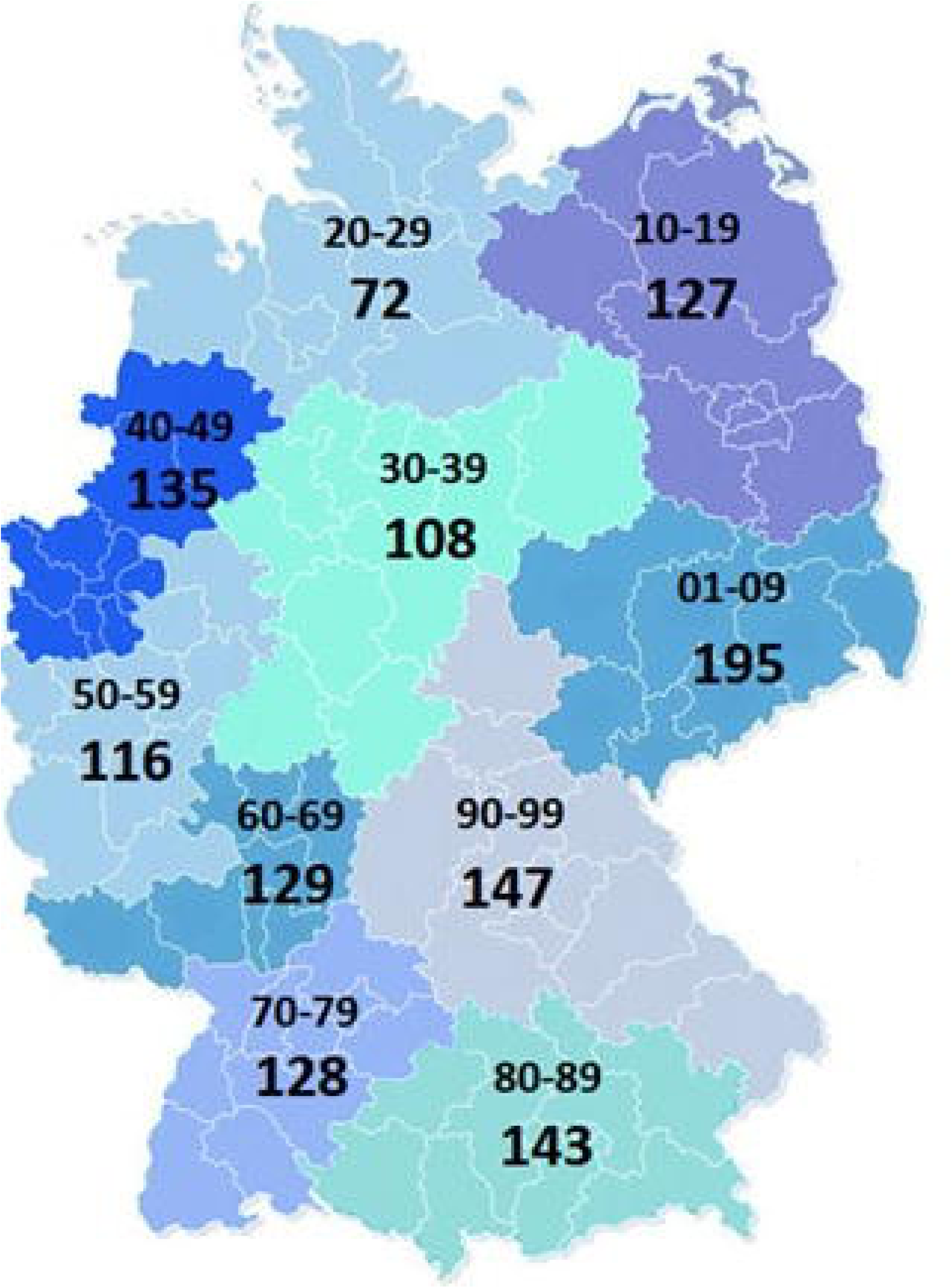
Mean incidence of COVID-19 infection per 100,000 inhabitants in Germany by the ZIP-code region beginning from the 2nd COVID-19-wave (calendar week 40^th^ 2020) until the end of the survey (3rd calendar week 2021) Source: Robert Koch-Institut: SurvStat@RKI 2.0, https://survstat.rki.de, Data request: 02.08.2021

### HCW status per facility type and specialty

Two thousand seventy medical facilities included in the analysis comprised a total of 25,113 HCW in the respective fields of specialisation (see Table 2). GIE private practices reported performing on average 21.2 (SD=15.3) procedures per day. In contrast, Non-GIE practices stated to carry out significantly more procedures 34.6 (SD=27.7, p<0.01). Clinics performed overall significantly more procedures compared to private practices (41.5 (SD=23.4) vs. 32.9 (SD=26.8), p<0.01), with Non-GIE performing significantly more procedures as GIE (58.6(SD=35.6) vs. 24.9 (SD=12.9), p <0.01).

**Table 2:**
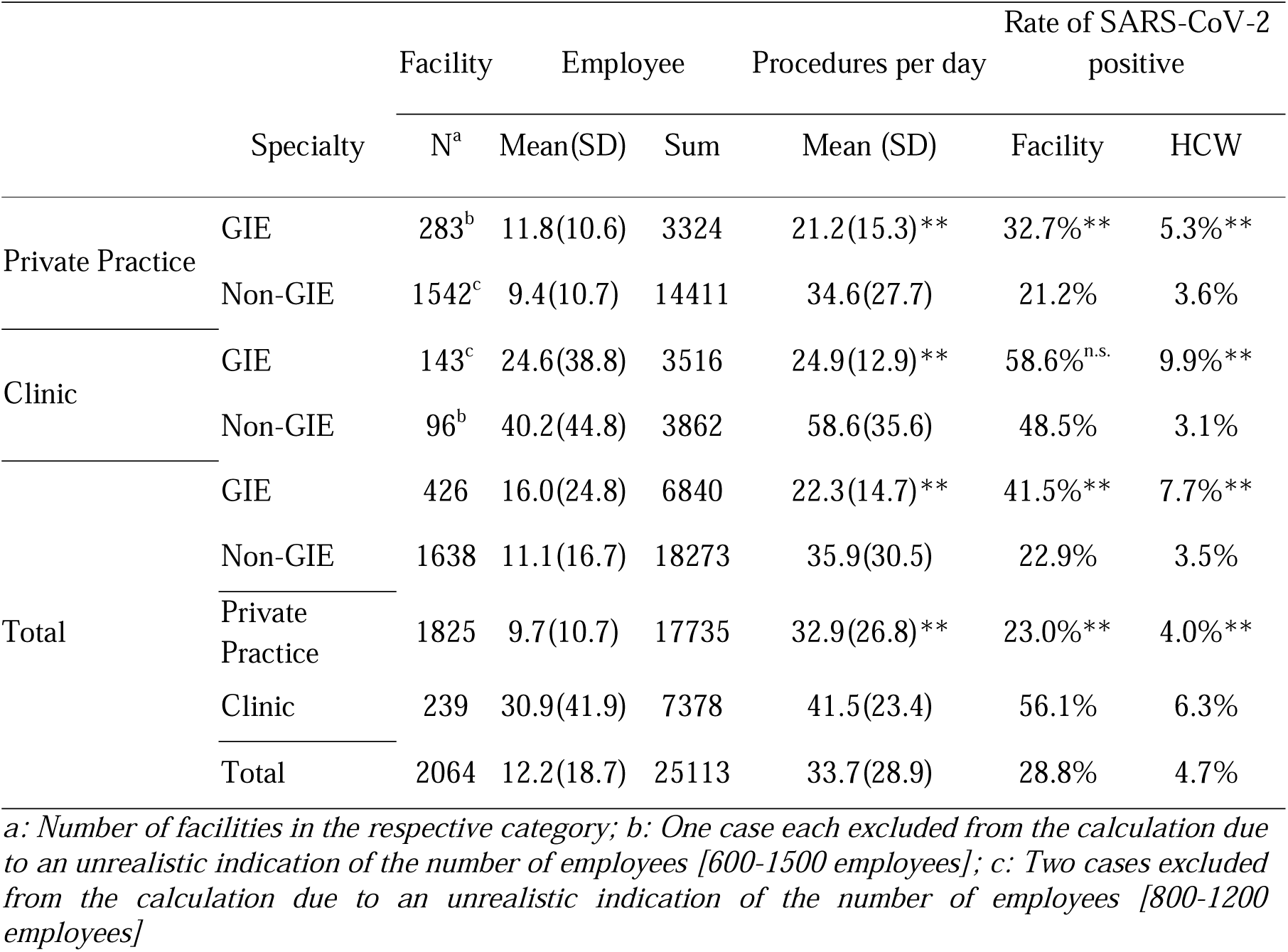
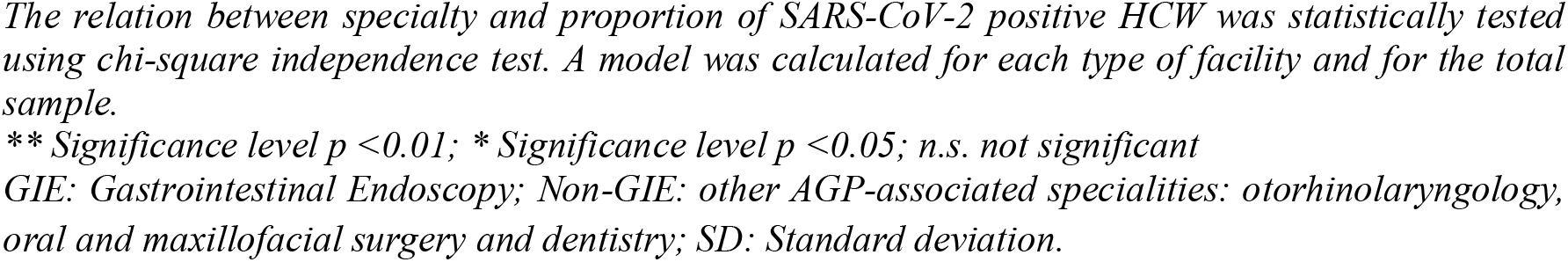
SARS-CoV-2-positive HCW by facility type, specialty, and collective size in the questionnaire

32.7% of the GIE private practices reported having had at least one COVID-19 case among HCW, whereas this share was significantly lower in Non-GIE (21.2%, p<0.01). The proportion of SARS-CoV-2 infections was significantly higher in the clinical setting than private practices in both specialties (56.1% vs. 23.0%, p<0.01) and accounted in GIE for 58.6% and 48.5% in Non-GIE. However, this difference was not significant. Overall, the rate of HCW who were reported to have had a COVID-19 infection was 4.7%. The rate was significantly higher in the GIE compared to the Non-GIE (7.7% vs. 3.5%, p>0.01). The number of infected HCW was significantly higher in clinics than in private practices (6.3% vs. 4.0%, p<0.01). A significant difference between the examined specialties was reported for private practices, with 5.3% of GIE HCW and only 3.6% SARS-CoV-2-infected HCW in Non-GIE facilities. The same applies to the clinical setting: a significantly higher proportion of SARS-CoV-2-positive employees was also found for GIE in comparison to Non-GIE (9.9% vs. 3.1%, p<0.01). Consequently, a higher risk of infection was reported for GIE than for the other AGP-associated disciplines for private practices and clinical settings.

### Source of infection

The implied source of infection identified by the heads of the facilities was predominantly the private environment (66%), followed by an unclear origin of infection (14%) (Figure 2). Only 14% of cases were attributed to patient contact or medical procedures. In GIE, the proportion of HCW with an implied source of infection during patient care (“During interventions” and “During other patient contacts”) was significantly higher than in Non-GIE 10% vs. 5%, p < 0.001, and 13% vs. 4%, p < 0.001 respectively). Accordingly, the proportion of employees who reported being infected in their private environment was substantially higher in Non-GIE specialty (73% vs. 56%, p < 0.001).

**Figure 2:**
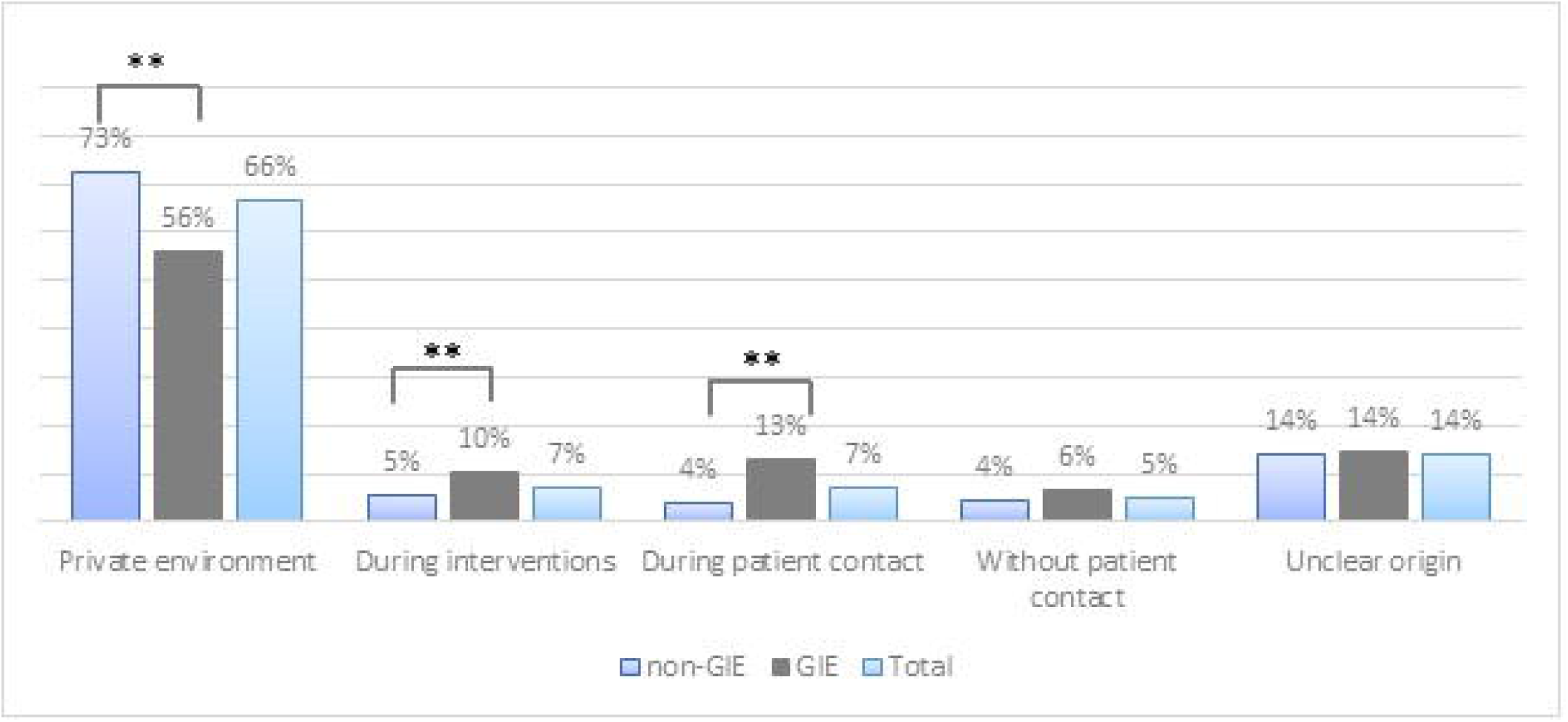
Implied source of infection by specialisation The confounding effect of the medical facility was adjusted using Cochran-Mantel-Haenszel test. One model was calculated for each implied source of infection. Only significant differences are marked. ** Significance level p <0.01; * Significance level p <0.05; n.s. not significant GIE: Gastrointestinal Endoscopy; Non-GIE: other AGP-associated specialties: otorhinolaryngology, oral and maxillofacial surgery, and dentistry.

### Pre-interventional testing

In private practices, Non-GIE specialties were testing their outpatients significantly more frequently than GIE (15.2% vs. 7.7%, p < 0.01). Regarding clinical settings, both out- and inpatients were significantly more often tested pre-interventionally compared to private practices. 58.9% of Non-GIE outpatients and 72.2% GIE outpatients were tested before procedures (p < 0.05), whereas among inpatients in all specialties, the testing rate was relatively high, accounting for 96.1% and 93.1% in Non-GIE and GIE, respectively. Furthermore, GIE clinics reported to test both their out-and in patients significantly more frequently using antigen than PCR (36.8% vs. 21.1%, p < 0.01 and 23.4% vs. 11.7% p<0.05 respectively) (Table 3).

**Table 3:**
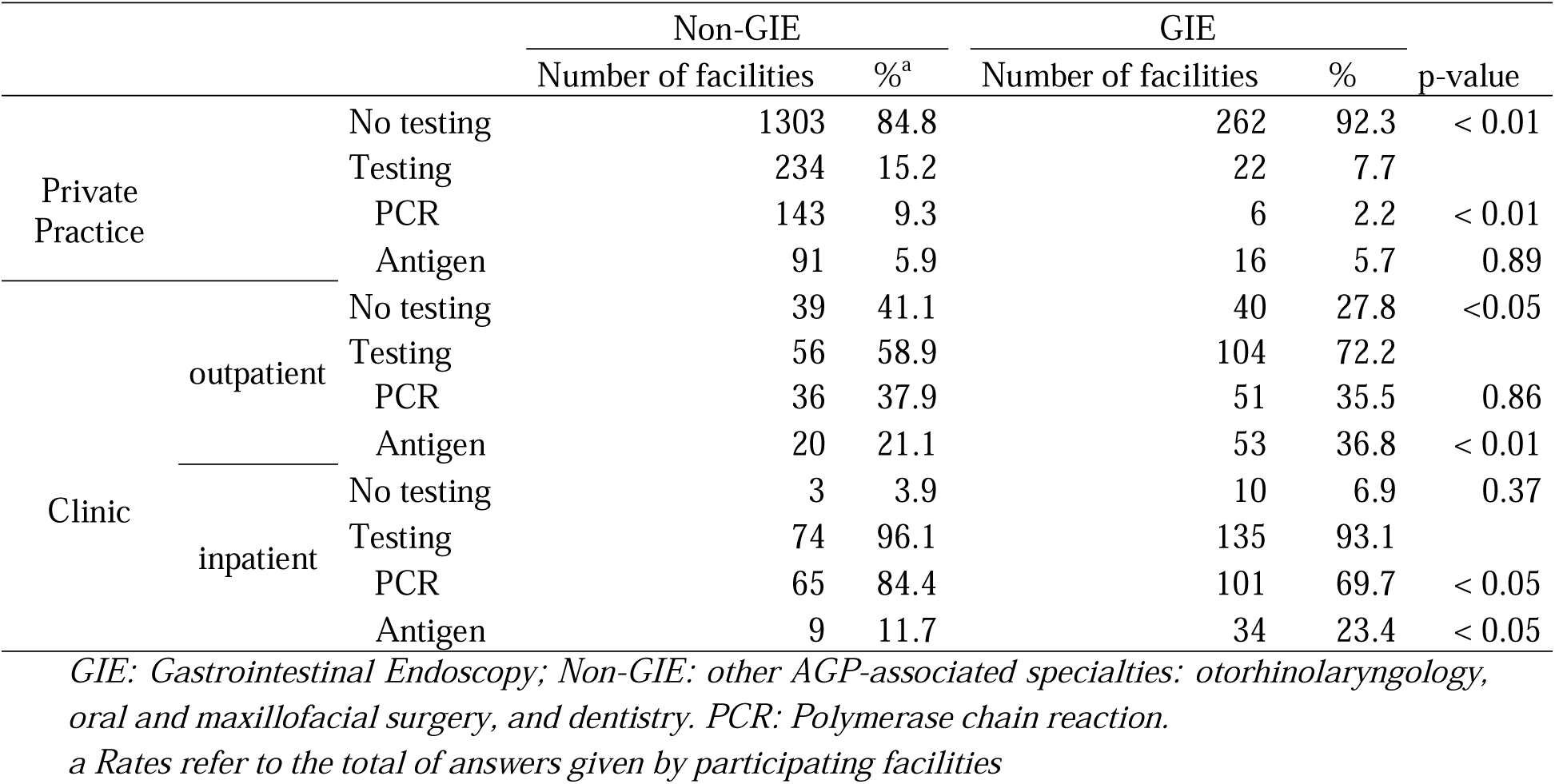
Pre-interventional testing by facility type and specialty

### Treatment of confirmed SARS-CoV-2 cases

In total, GIE medical facilities have treated more often confirmed SARS-CoV-2 cases (32.4% vs. 24.7%, p <0.01). GIE private practices stated to have treated confirmed SARS-CoV-2 patients in 7.7% of cases being significantly lower than Non-GIE 21.8% (p<0.01). On the contrary, GIE clinics treated more SARS-CoV-2 confirmed cases than Non-GIE (80.7% vs. 72.2%, p=0.548). Overall, clinics treated confirmed SARS-CoV-2 patients almost four times more often than private practices (77.3% vs. 19.5%, p < 0.01) (Table 4).

**Table 4:** Treated confirmed SARS-CoV-2 cases by facility type and specialty

|  | Non-GIE |  | GIE |  | Total |  | p-value |
| --- | --- | --- | --- | --- | --- | --- | --- |
|  | Number of facilities | % | Number of facilities | % | Number of facilities | % |  |
| Private practice | 336 | 21.8 | 22 | 7.7 | 356 | 19.5 | <0.01 |
| Clinic | 70 | 72.2 | 117 | 80.7 | 187 | 77.3 | 0.548 |
| Total | 406 | 24.7 | 139 | 32.4 | 545 | 26.3 | <0.01 |

*GIE: Gastrointestinal Endoscopy; Non-GIE: other AGP-associated specialties: otorhinolaryngology, oral and maxillofacial surgery, and dentistry.*

### Multivariate analysis of the risk factors

Among examined factors, several ZIP-code regions were significantly associated with HCW infection in medical facilities. As ZIP-region 20-29 having the lowest incidence within examined period in time was chosen as a reference group, multivariate analysis revealed, consistently, odds ration corresponding to increased risk of SARS-Cov-2 infectio associated with nine other/remaining ZIP-regions. However, not all of this associations were significant. Particularly, regions 60 to 89, bordering Austria, Switzerland, and France, were significantly associated with an two-to three-fold increased risk of infection among HCW. And in the ZIP-code region 01-09, having the highest incidence in Germany, the risk of infection was 2.04 time higher compared to the reference group (95% CI 1.124-3.689, p=0.019). Considering further influencing variables, in the clinical setting there was 2.63 times (95% CI 2.504 - 2.817, p <0.01) increased risk of infection. GIE was also significantly associated with 2.35 times (95% CI 2.245-2.498, p<0.01) increased risk of SARS-CoV-2 infection among HCW. The number of procedures carried out per day in a medical facility increased the probability of infection, however, only marginally (OR 1.01, 95% CI 1.007-1.014), p<0.01). Having not treated any confirmed SARS-CoV-2 patients was related to a lower risk of contagion (OR 0.72, 95% CI 0.507-1.025, p=0.068); however, this association was marginally insignificant. Pre-interventional testing has not significantly influenced the occurrence of the SARS-CoV-2 infection among HCW (Table 5).

**Table 5:** Summary of the risk factors associated with SARS-CoV-2 infections among HCW

| Category | Dimension | OR | CI 95% |  | p-value |
| --- | --- | --- | --- | --- | --- |
|  |  |  | LB | UB |  |
| ZIP-code region | 01-09 | 2.04 | 1.124 | 3.689 | 0.019 |
|  | 10-19 | 1.86 | 1.063 | 3.256 | 0.03 |
|  | 30-39 | 1.18 | 0.700 | 1.987 | 0.535 |
|  | 40-49 | 1.46 | 0.880 | 2.423 | 0.143 |
|  | 50-59 | 1.41 | 0.859 | 2.324 | 0.174 |
|  | 60-69 | 3.31 | 2.071 | 5.296 | <0.01 |
|  | 70-79 | 2.34 | 1.449 | 3.763 | <0.01 |
|  | 80-89 | 2.51 | 1.594 | 3.955 | <0.01 |
|  | 90-99 | 1.11 | 0.663 | 1.872 | 0.683 |
|  | 20-29 [RG] |  |  |  |  |
| Healthcare delivery setting | clinics | 2.63 | 2.504 | 2.817 | <0.01 |
|  | private practices [RG] |  |  |  |  |
| Medical specialty | GIE | 2.35 | 2.245 | 2.498 | <0.01 |
|  | Non-GIE [RG] |  |  |  |  |
| Procedures p. day |  | 1.01 | 1.007 | 1.014 | <0.01 |
| Confirmed SARS-CoV-2 cases | no | 0.72 | 0.507 | 1.025 | 0.068 |
|  | yes |  |  |  |  |
| Pre-interventional testing | PCR | 0.98 | 0.678 | 1.423 | 0.659 |
|  | Antigen | 1.11 | 0.741 | 1.655 | 0.397 |
|  | No testing [RG] |  |  |  |  |

HCW: healthcare worker; GIE: Gastrointestinal Endoscopy; Non-GIE: other AGP-associated
specialties: otorhinolaryngology, oral and maxillofacial surgery, and dentistry
OR: odds ratio, CI: confidence interval, LB: lower bound, UB: upper bound, RG reference group

## DISCUSSION

This study is the first to present cumulated data on the prevalence of SARS-CoV-2 infection in HCW in different medical subspecialties and healthcare delivery settings. In particular, this manuscript focuses on medical disciplines associated with AGP, including GIE, ORL, OMS, and dentistry. Data from private practices and clinics were collected via a nationwide questionnaire-based survey conducted in Germany (83.02 Mio inhabitants) between the 16th of December and the 24th of January,[7].

Our study shows a nationwide total HCW-infection rate accounting for 4.7% in the four examined specialties. Due to the current state of research, there is no consensus regarding the increased infection rate of HCW compared to the normal population so far. For instance, Jungo et al. (2021) could not confirm an increased rate of infections in dental offices as compared to the normal population,[8], whereas, on the contrary, in a large Danish cohort, an increased risk for HCW could be shown based on the seroprevalence compared to blood donors,[4]. RKI officially reported 2,134,936 confirmed cases for Germany on 01/24/2021 by the end of the survey,[9]. According to this data, approximately 2.6% of the German population had been infected with SARS-CoV-2 by the time of the study. Based on this insight, it could be inferred that HCW involved in AGP have an increased risk of infection compared to the general population. However, it should be taken into account that not all infections are recorded in official registries due to several reasons, such as a high rate of asymptomatic or mild courses of Covid-19,[10, 11]. For instance, the project “Dunkelzifferradar,” funded by the Federal Ministry of Education and Research, uses a mathematical model to estimate unreported cases of SARS-CoV-2 infections, assuming a calculation of approximately 6.5 million infected people by 01/24/2021. This would result in a Germany-wide prevalence of 7.8% at the time of the survey,[12, 13]. Another estimate is given by the Gutenberg Covid-19 study, which indicates that around 42% of infections in Germany are not detected, resulting in a Germany-wide prevalence of 4.5% at the time of the survey,[14]. Considering these estimates, an increased risk for SARS-CoV-2 infection for HCW in the examined disciplines compared to the general population cannot be clearly demonstrated by our study, with the exception of HCW in a GIE clinical setting with a prevalence of SARS-CoV-2 infections of 9.9%. Nevertheless, data on the prevalence of SARS-CoV-2 infections among HCW are inconsistent, ranging from 4.3% to 32.5% in the current scientific literature,[3, 4, 15-21]. It seems obvious that the prevalence in HCW depends on multiple factors, such as, healthcare delivery setting, local infection occurrence during the investigation period, pre-interventional testing, political and social measures.

Our study revealed a significantly higher proportion of infected HCW in clinics with 6.3% compared to 4.0% in private practices. Furthermore, according to the multivariate model, clinical setting was associated with more than doubled risk of SARS-CoV-2 infection among HCW. Assuming that patients are a potential source of infection in a medical facility, the number of patients seen per day and, thus, procedures performed might influence the risk of infection. This consideration was confirmed in our study by a significant association of occurrence of infection and number of procedures performed per day. Indeed, clinics perform on average more procedures than private practices, bringing HCW at higher risk of transmission. Furthermore, besides the higher accumulation of people, clinics treat more patients with urgent or emergency procedures. In line with this, clinics have treated confirmed SARS-CoV-2 patients almost four times more often than private practices. According to the multivariate model, treatment of confirmed SARS-CoV-2 cases was tending to be a risk of infection in a medical unit, however, this association was marginally not significant.

In our study, GIE was shown to have a significantly higher positive HCW rate than Non-GIE in both examined healthcare delivery settings. Interestingly, GIE clinics have been stated to treat confirmed SARS-CoV-2 cases more often than Non-GIE, whereas in private practices, an opposite tendency was observed: GIE reported having had significantly fewer confirmed cases under treatment. Moreover, GIE performed significantly fewer procedures per day compared to Non-GIE medical disciplines. Despite that, GIE showed a significantly higher HCW infection rate in both healthcare delivery settings. The reason for the higher infection rates in the GIE, specifically in a clinical setting, might be the higher rate of non-elective procedures conducted on COVID-19 patients,[22]. Furthermore, Repici et al. (2020) discussed other specific characteristics of GIE applicable to private practices, such as the high level of unnoticed exposure of HCW during endoscopic procedures,[23, 24]. Many COVID-19 patients show gastrointestinal symptoms,[25]; hence they might undergo endoscopic examination before the identification of SARS-CoV-2 infection. Furthermore, COVID-19 patients often require endoscopic procedures such as bronchoscopies due to pulmonary involvement and gastroscopies during intensive care stays in case of bleeding complications,[26]. Another reason for the higher rate of SARS-CoV-2 positive HCW in GIE in clinics might be shifting GIE HCW into COVID-19 wards implicating direct contact to confirmed COVID-19 patients. Data on the risk of infection among HCW in designated Covid-19 wards is heterogeneous,[27-29]. A monocentric survey in a tertiary care hospital in Turkey showed an increased risk of infection for HCW working on COVID-19 wards compared with Non-COVID-19 areas. However, incorrect handling and non-compliance with the hygienic/distance rules among HCW were risk factors associated with infection,[3].

One possible action to prevent transmission of SARS-CoV-2 into medical facilities is pre-interventional testing. According to the findings of our study, in private practices, pre-interventional testing of patients was performed only in roughly 10% of the cases, with Non-GIE testing twice as often as GIE. In the clinical setting, all specialties tested their patients before intervention substantially more frequently, with inpatients being tested in over 90% of cases. Despite that, the prevalence of SARS-CoV-2 positive HCW was significantly higher in clinics than private practices, indicating that testing may not play a crucial role at low to moderate incidence levels as discussed by guidelines,[30, 31]. In line with that insight, multivariate model revealed no significant association of pre-interventional testing with occurrence of the SARS-Cov-2 infection in a medical unit. On the one hand, it suggests that AGP can be safely performed by HCW using adequate personal protective equipment and following hygienic concepts,[32]. On the other hand, it raises the question of how COVID-19 cases invade medical facilities despite a high rate of pre-interventional testing, especially in clinics. Considering the relatively high proportion of antigen tests used as SARS-CoV-2 detection tools, this might indicate poor sensitivity of these tests, accounting, according to Kahn et al. (2021), for about 50-60%,[33].

The prevalence of SARS-CoV-2-infected HCW might strongly depend on the local occurrence of infection. According to the multivariate model ZIP-code regions having higher mean incidence within examined period of time were associated with increased risk of infection in the medical facilities compared to the ZIP-region 20-29, having the lowest observed mean incidence. However not all associations were significant. For instance, the ZIP-region 90-99 having the second-highest mean incidence in the considered period was not significantly different from the region with the lowest mean incidence. This observation highlights the difficulty of associating the SARS-CoV-2 incidence to the prevalence of SARS-CoV-2 infections in medical facilities. Firstly, a mean incidence reflects a tendency across a defined period neglecting development over time. Secondly, medical facilities might apply protective measures and guidelines or cancel procedures and limit access to the facility for non-patients to prevent transmission of the infection. Thirdly political and social measures could be taken and differ even between counties and districts.

Finally, even assuming HCW to have higher occupation-related infection risk than other professions, infection outside the workplace cannot be ruled out. Thus, preliminary results of a study from Italy suggest local infections in the private environment rather than occupational exposure,[34]. This is in line with the results of our study, whereas the primary source of infection stated was the private environment in all questioned specialties. Nevertheless, the share of COVID-19 infection attributed to the workplace was still significantly higher in GIE facilities. In particular, in GIE it is significantly more often associated with interventions/procedures or other patient contacts. This underlines the increased risk for HCW in GIE.

Like other cross-sectional studies, our study has some limitations. Due to the recruitment strategy via the professional associations, a selection bias cannot be ruled out. In particular, facilities that established elaborate protection and hygiene measures might have been higher motivated to participate. On the other hand, facilities with infected HCWs may also be more motivated to participate. Moreover, there is an uneven distribution of the medical facility types between examined specialties. For instance, in dental medicine, hardly any clinic was represented in comparison to the more than 1000 participating private practices.Nonetheless, Non-GIE specialties had significantly more private practices due to the regional specificity of the respective fields of activity. Another shortcoming of the study worth mentioning is that this study is cross-sectional inquiring information over a considerable period comprising three quarters of the year 2020. Moreover, all calculations presented in the manuscript are based on the assessments and judgments made for a private practice or a hospital ward and its workforce by one person.

Despite the limitation mentioned above, the present study is the first to provide data on prevalence and revealing risk factors of SARS-CoV-2 infection among HCW in medical disciplines associated with AGP, such as GIE, ORL, OMS, and dentistry. Due to the results provided in this scientific manuscript, GIE seems to be at a higher risk of infection compared to the other investigated disciplines.

## Data Availability

Data are available upon reasonable request.

## ACKNOWLEDGMENTS

We thank all medical facilities for participating in this study.

## CONTRIBUTORS

Conception and design: CR, AE, HM, MK, SZ, CTH. Analysis and interpretation of the data: AM, CR, AE, HM, MK, SZ. Drafting of the article: CR, AM, SZ, MK. Critical revision of the article for important intellectual content: AE, HM. Final approval of the article: all authors; Statistical expertise: AM. Administrative, technical or logistic support: GH, CB, AB, JR, TS, JZ, BA, RF, JH, CA, FL. Collection and assembly of data: CR, AE, MK, SZ.

## ETHICS APPROVAL

The study was conducted in accordance with the Declaration of Helsinki and Good Clinical Practices. An approval of the study was given by the Ethics Committee of the University Hospital of Augburg (713/20 S-SR).

## PATIENT CONSENT FOR PUBLICATION

Not required.

## DATA AVAILABILITY STATEMENT

Data are available upon reasonable request.

