## Supplementary material for "Health-care workers in gastrointestinal endoscopy are at higher risk for SARS-CoV-2 infection compared to other aerosol-generating disciplines"

**Supplemental material**

*S1: Absolute and percentage distribution of cases by type of institution and speciality*

|  | Private Practice | |  | Clinic | |  |
| --- | --- | --- | --- | --- | --- | --- |
| Specialisation | Number of  Facilities | % of all facilities |  | Number of  Facilities | % of all facilities | Total |
| GIE | 284 | 13.5% |  | 145 | 6.9% | 429 |
| ORL | 395 | 18.8% |  | 66 | 3.1% | 461 |
| Dentistry | 1094 | 52.1% |  | 20 | 1.0% | 1114 |
| OMS | 55 | 2.6% |  | 11 | 0.5% | 66 |
| Others^a^ | 10 | 0.5% |  | 16 | 0.8% | 26 |
| Total | 1838 | 87.7% |  | 258 | 13.3% | 2096 |

S2: Absolute and percentage distribution of participants by ZIP-codes and its corresponding incidence

|  | N | % | Mean incidence | SD |
| --- | --- | --- | --- | --- |
| 01-09 | 100 | 4.83% | 195 | 143.0 |
| 10-19 | 131 | 6.33% | 127 | 73.4 |
| 20-29 | 253 | 12.22% | 72 | 38.3 |
| 30-39 | 207 | 10.00% | 108 | 60.6 |
| 40-49 | 210 | 10.14% | 135 | 56.3 |
| 50-59 | 231 | 11.16% | 116 | 53.0 |
| 60-69 | 215 | 10.39% | 129 | 66.7 |
| 70-79 | 230 | 11.11% | 128 | 61.9 |
| 80-89 | 278 | 13.43% | 143 | 71.7 |
| 90-99 | 215 | 10.39% | 147 | 90.3 |
| Total | 2070 | 100.00% |  |  |

epici A, Pace F, Gabbiadini R et al. Endoscopy units and the

COVID-19 outbreak: a multi-center experience from Italy. Gastroen-

terology. 2020; 159(1): 363–66e3

Repici A, Pace F, Gabbiadini R et al. Endoscopy units and the

COVID-19 outbreak: a multi-center experience from Italy. Gastroen-

terology. 2020; 159(1): 363–66e3

Repici A, Pace F, Gabbiadini R et al. Endoscopy units and the

COVID-19 outbreak: a multi-center experience from Italy. Gastroen-

terology. 2020; 159(1): 363–66e3
